# Recurrent respiratory viral diseases and chronic sequelae due to dominant negative IFIH1

**DOI:** 10.1101/2020.07.01.20105379

**Authors:** Christin L. Deal, Timothy J. Thauland, Rebecca Signer, Stanley F. Nelson, Undiagnosed Diseases Network, Hane Lee, Manish J. Butte

## Abstract

Viral respiratory infections are the most common childhood infection worldwide. However, even common pathogens can have significant consequences in the context of patients with primary immunodeficiency diseases. More than half or viral infections annually are due to rhinovirus/enterovirus strains. Most clinical manifestations of viral infection are mild. However 3% of cases result in hospitalization in patients who have no other known risk factors. These patients may have an inborn error of immunity, a genetic susceptibility to viral infections. Here we present the case of an adult male who suffered respiratory viral infections his whole life and developed chronic, inflammatory damage to sinuses and lungs as a consequence. Genomic sequencing identified compound heterozygous variants in the IFIH1 gene, encoding the protein Melanoma Differentiation Association Protein 5 (MDA5), a RIG-I-like cytoplasmic sensor of RNA intracellular infections. We show a dominant negative effect on these variants on the level of interferon-induced expression of MDA5 protein. This work supports that loss-of-function variants in IFIH1 affect the sensing of viral infections. Underlying genomic variants may dictate the point at which recurrent, respiratory viral infections leave commonplace experience and incur lasting damage.

## To the Editor

Viral respiratory infections are the most common childhood infection worldwide. However, even prevalent pathogens can have significant consequences in patients with primary immunodeficiency diseases (PID). More than half of viral infections annually are due to rhinovirus/enterovirus strains. Most clinical manifestations of viral infections are mild. However, 3% of cases result in hospitalization in patients who have no other known risk factors (1). These patients may have a genetic susceptibility to viral infections.

Interferon-induced helicase C domain-containing protein 1 (*IFIH1*) is a gene encoding Melanoma Differentiation Association Protein 5 (MDA5), a RIG-I-like cytoplasmic sensor of long double-stranded RNA, which is key in innate immune recognition of RNA viruses. On binding viral dsRNA, MDA5 triggers signaling molecules IPS-1, IRF3 and IRF7 resulting in transcription of type 1 interferons (2). Autosomal dominant gain-of-function variants in *IFIH1* are known to cause Aicardi-Goutières syndrome (3). Other variants have been found to predispose to autoimmune conditions like type-1 diabetes, vitiligo, autoimmune thyroiditis, and SLE (4). Furthermore, biallelic loss of function variants in *IFIH1* result in severe infections due to common viral illnesses (2,3), and one study suggested a possible dominant negative role for heterozygous loss-of-function variants (1).

Patients with loss-of-function variants in *IFIH1* are susceptible to common viral pathogens, particularly human rhinovirus. Importantly, patients with these variants have a normal evaluation of their circulating immune cells, because the defect does not affect the generation or function of adaptive immune cells like T and B cells. However, patients carrying these variants are at risk of severe clinical manifestations due to viral infections as well as bacterial superinfections. Understanding who is susceptible can allow for better preventative care for these individuals.

## Case Presentation

A 57-year-old male was referred to the Undiagnosed Diseases Network for chronic pansinusitis. Throughout childhood he suffered numerous viral upper respiratory infections that were often accompanied by wheezing. In his late 20s he developed wheezing episodes with shortness of breath that were refractory to all treatments other than oral corticosteroids. In his 30s and 40s these episodes became more frequent, increasing from two episodes annually to five episodes annually. In his 50s he developed anosmia and was found to have nasal polyps. He underwent 3 sinus surgeries for polyposis and sinusitis as well as tympanostomy tubes bilaterally for bilateral ear pain. After surgical intervention, he continued to suffer from pansinusitis **(Figure 1A)**. He did not suffer from fungal infections, skin infections, invasive bacterial infections, or chronic viral infections such as warts, CMV, or EBV.

**Figure 1.**
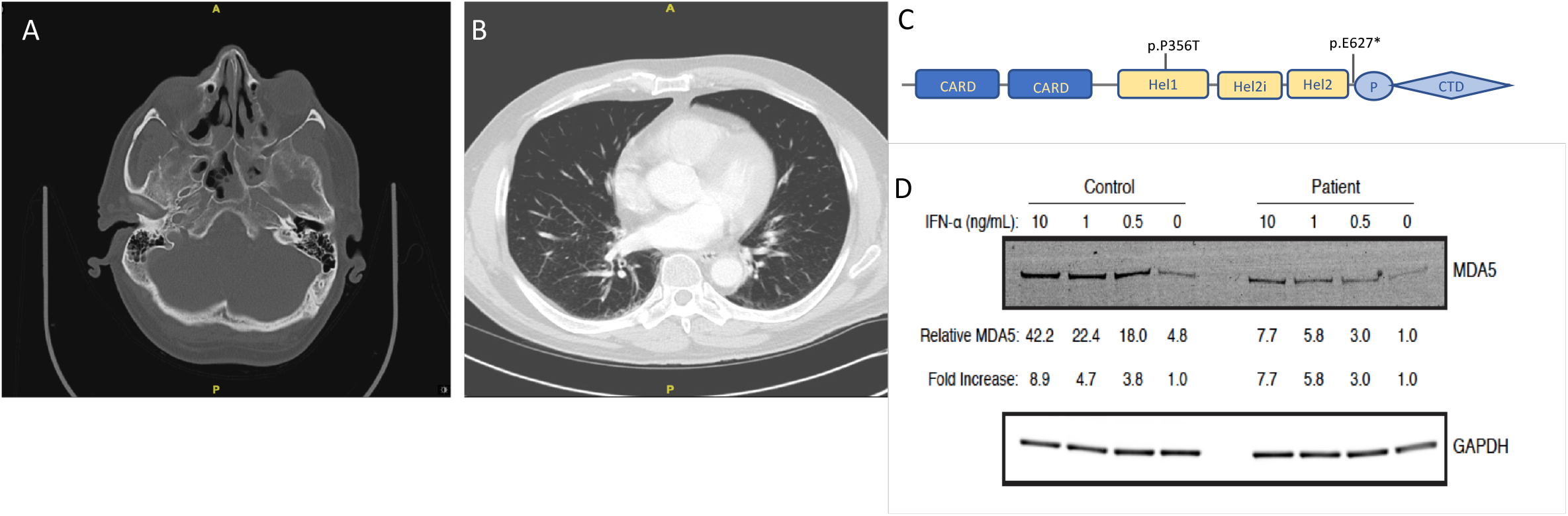
**(A)** Circumferential mucosal thickening in the bilateral maxillary sinuses and sphenoid sinuses. Near-complete opacification of the scattered ethmoid air cells. The sphenoid ethmoidal and frontoethmoidal recesses are opacified bilaterally. **(B)** Areas of ground glass opacification, left greater than right, reside predominantly in the lower lobes of the lung which are likely due to atelectasis. In addition, the right lower lobe has mild bronchiectasis. **(C)** IFIH1 Domains. Our patient’s nonsense mutation results in termination of the protein prior to the critical C-terminal regulatory domain which is critical for recognizing and binding viral dsRNA. **(D)** PBMC from a healthy control and the patient were stimulated overnight with the indicated concentrations of IFNα. The amounts of MDA5 relative to the unstimulated patient cells are shown. Band intensities were normalized to the GAPDH loading control. Fold increase in MDA5 upon IFNα stimulation are also shown for the healthy control and patient. Results are representative of two independent experiments.

He had eosinophilic asthma but normal total IgE. He had negative skin prick tests to aeroallergens and undetectable serum-specific IgE to aeroallergens. He tolerates aspirin. His chest CT showed evidence of atelectasis and bronchiectasis **(Figure 1B)**. Immune evaluation revealed mildly low total IgG of 577 mg/dL due to mildly low IgG1 and IgG3 subclasses. He had normal lymphocyte subsets, normal pneumococcal titers after immunization with PPV23 (pneumovax), and protective tetanus and diphtheria titers long after immunization **(Table 1)**.

**Table 1.**
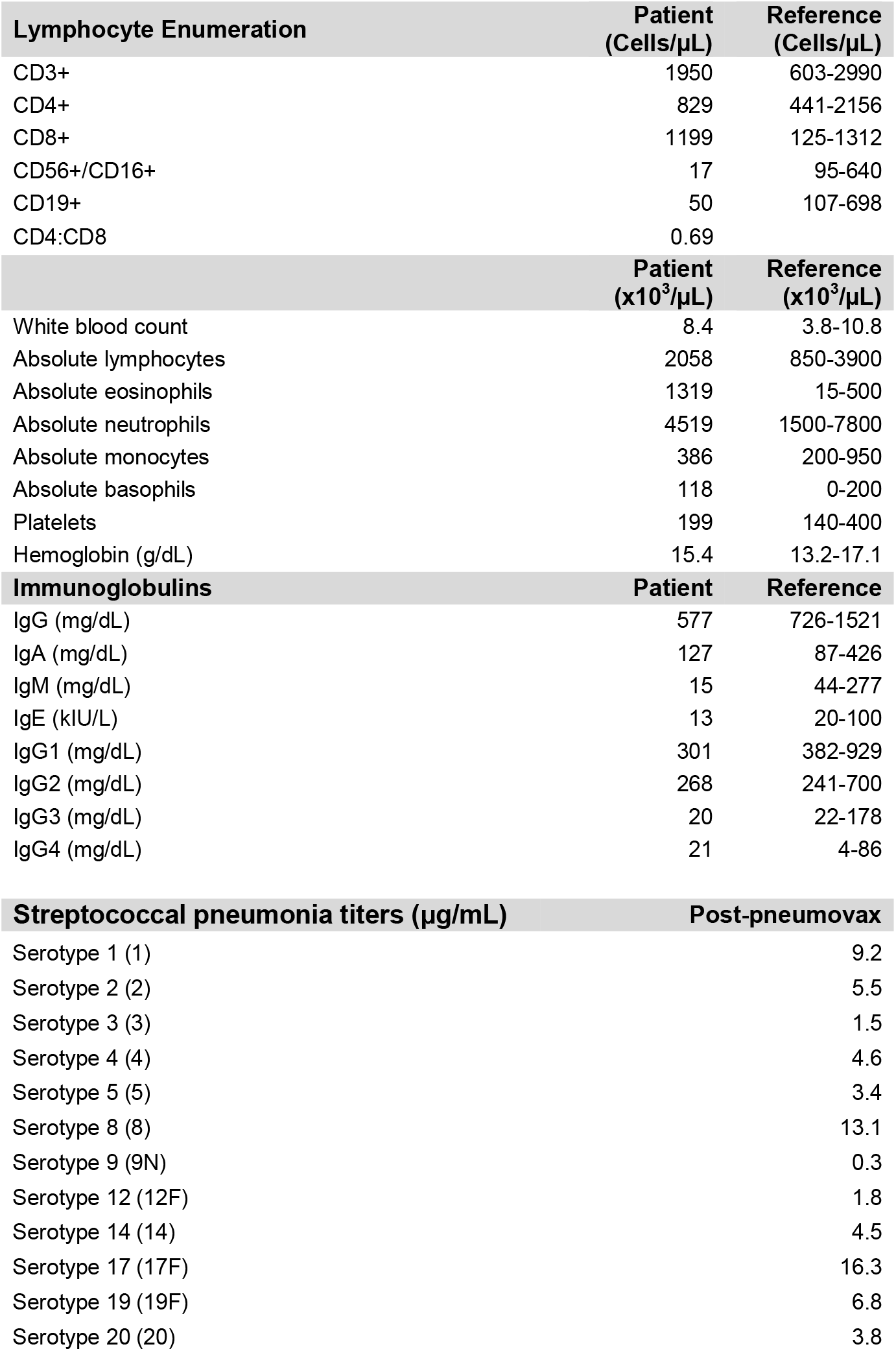
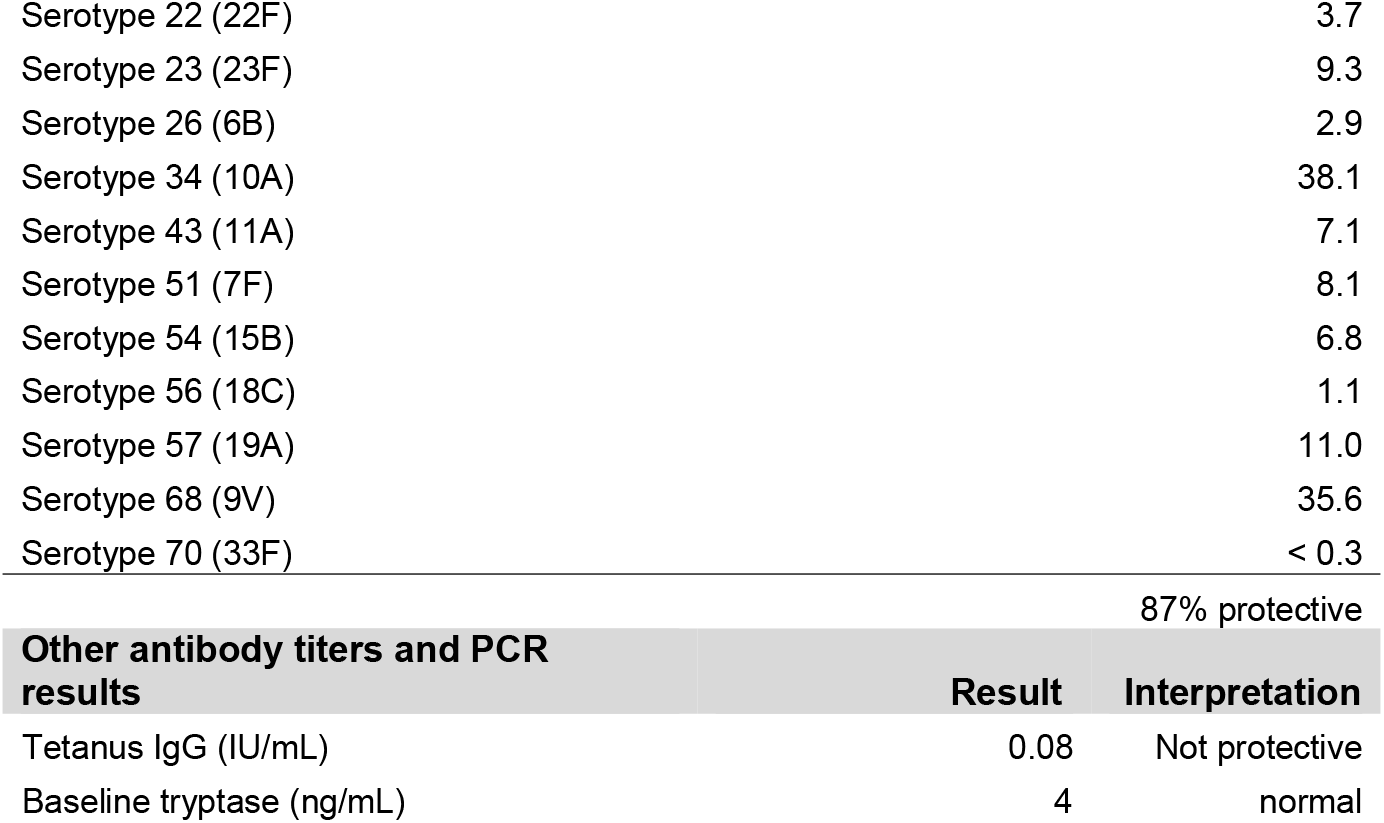
Summary of immune evaluation.

| **Lymphocyte Enumeration** | **Patient**<br>**(Cells/μL)** | **Reference**<br>**(Cells/μL)** |
| --- | --- | --- |
| CD3+ | 1950 | 603-2990 |
| CD4+ | 829 | 441-2156 |
| CD8+ | 1199 | 125-1312 |
| CD56+/CD16+ | 17 | 95-640 |
| CD19+ | 50 | 107-698 |
| CD4:CD8 | 0.69 |  |
|  | **Patient**<br>**($\times 10^3$/μL)** | **Reference**<br>**($\times 10^3$/μL)** |
| White blood count | 8.4 | 3.8-10.8 |
| Absolute lymphocytes | 2058 | 850-3900 |
| Absolute eosinophils | 1319 | 15-500 |
| Absolute neutrophils | 4519 | 1500-7800 |
| Absolute monocytes | 386 | 200-950 |
| Absolute basophils | 118 | 0-200 |
| Platelets | 199 | 140-400 |
| Hemoglobin (g/dL) | 15.4 | 13.2-17.1 |
| **Immunoglobulins** | **Patient** | **Reference** |
| IgG (mg/dL) | 577 | 726-1521 |
| IgA (mg/dL) | 127 | 87-426 |
| IgM (mg/dL) | 15 | 44-277 |
| IgE (kIU/L) | 13 | 20-100 |
| IgG1 (mg/dL) | 301 | 382-929 |
| IgG2 (mg/dL) | 268 | 241-700 |
| IgG3 (mg/dL) | 20 | 22-178 |
| IgG4 (mg/dL) | 21 | 4-86 |
| **Streptococcal pneumonia titers (μg/mL)** | **Post-pneumovax** |  |
| Serotype 1 (1) | 9.2 |  |
| Serotype 2 (2) | 5.5 |  |
| Serotype 3 (3) | 1.5 |  |
| Serotype 4 (4) | 4.6 |  |
| Serotype 5 (5) | 3.4 |  |
| Serotype 8 (8) | 13.1 |  |
| Serotype 9 (9N) | 0.3 |  |
| Serotype 12 (12F) | 1.8 |  |
| Serotype 14 (14) | 4.5 |  |
| Serotype 17 (17F) | 16.3 |  |
| Serotype 19 (19F) | 6.8 |  |
| Serotype 20 (20) | 3.8 |  |

|  |  |
| --- | --- |
| Serotype 22 (22F) | 3.7 |
| Serotype 23 (23F) | 9.3 |
| Serotype 26 (6B) | 2.9 |
| Serotype 34 (10A) | 38.1 |
| Serotype 43 (11A) | 7.1 |
| Serotype 51 (7F) | 8.1 |
| Serotype 54 (15B) | 6.8 |
| Serotype 56 (18C) | 1.1 |
| Serotype 57 (19A) | 11.0 |
| Serotype 68 (9V) | 35.6 |
| Serotype 70 (33F) | < 0.3 |

87% protective
| Other antibody titers and PCR results |  |  |
| --- | --- | --- |
|  | Result | Interpretation |
| Tetanus IgG (IU/mL) | 0.08 | Not protective |
| Baseline tryptase (ng/mL) | 4 | normal |

He had a history of laparoscopic Nissen fundoplication for GERD. He was found to have an ascending aortic aneurysm, and family history was only remarkable for cardiac disease in mother, sister and brother. He recently developed signs of peripheral neuropathy. A part of his evaluation included ophthalmology screen which revealed ocular albinism **(Figure S1)**.

Whole exome sequencing revealed compound heterozygous variants in *IFIH1* resulting in NM_022168:c.1879G>T NP_071451.2:p.E627* (rs35744605) and c.1066C>A p.P356T (rs150317197) (5–8) **(Figure 1C**). The global minor allele frequency of the first variant in the population is roughly 0.003 and the second is 0.0006, which each fall below the population maximum allele frequency and so could be causative variants (9).

We pursued these variants knowing that a dominant negative variant could play a role in pathogenesis. The second variant has been biochemically studied in isolation, expresses well, and showed a near-normal ability to drive IFN-β in response to poly-I:C stimulation (3). The first variant, on the other hand, was ascribed to act in a dominant negative fashion when co-overexpressed in 293T cells with WT MDA5 protein (1), but this effect was not shown in primary cells. Thus, we sought to determine the amount of MDA5 in our patients’ cells. Since MDA5 expression is responsive to interferon α (IFN-α) stimulation, PBMCs from patient and control were stimulated overnight with IFN-α. Cells were lysed and examined by Western blot using a monoclonal antibody targeted to a region more C-terminal than the missense variant and more N-terminal than the truncation variant. In this way, the expression of both the truncated and mutated forms of the protein could potentially be detectable. We saw none of the truncated form of the protein in primary cells. The patient’s MDA5 levels were lower by over half at baseline as compared to control, when normalized to GAPDH. An increase in expression of MDA5 was seen after stimulation with IFN-α in both the subject and a control **(Figure 1D and Figure S2)**. In each condition, the relative amount of MDA5 expression was ∼20% of the control, well below the 50% one would expect if the protein amount were determined by the missense variant alone. These results show that the nonsense variant disproportionately and dominantly reduces the expression or stability of the protein bearing the missense variant.

## Discussion

There have been few case reports of *IFIH1* loss-of-function in children, and no cases yet reported in adults. Therefore, the complete clinical phenotype still remains poorly understood. Patients described thus far have a narrow spectrum of severe and recurrent respiratory viral infections with bacterial superinfections, but no other major viral infections (EBV, warts) (3). Prior cases report normal immune evaluation including responses to vaccines, with perhaps the exception of minor abnormalities in IgG and IgG subclasses, as seen here. Our patient was incidentally found to have ocular albinism and ascending aortic aneurysm. It is unlikely these findings are related to his *IFIH1* variants, but are pertinent findings in a poorly described phenotype.

The nonsense mutation in the middle of the protein eliminates the C-terminal regulatory domain that is essential for recognizing viral dsRNA. This results in a disruption of overall protein stability by the truncated variant (1). Therefore, in our patient, these variants resulted in decreased MDA5 expression. Our case thus supports the previous demonstration of a potential dominant negative mechanism in this disease.

The *IFIH1* gene tolerates a higher frequency of loss-of-function variants than would be expected for a rare disease. Evolutionary pressures on genes required for survival should, over time, “purify” away loss-of-function variants to low frequencies. On the other hand, loss-of-function variants of *IFIH1* carry protection from autoimmunity (8,10); conversely, gain-of-function variants with improved viral sensing predispose patients to autoimmunity (4). Thus, there may be greater tolerance to loss-of-function mutations than is seen in other PID genes, a potential example of “heterozygote advantage.” This reason may explain the relatively high frequency of nonsense variants in the population (0.3% for the nonsense variant, with an allele frequency highest in European populations per gnomAD). It is the delicate balance between an efficient immune response and autoimmunity that balances the selective pressures on this gene.

The phenotype of “susceptibility” to respiratory viral infections that have almost universal prevalence raises an important question about when it would be pragmatically useful to actually proceed with an immunological workup for individuals suffering from viral infections. We suggest that all patients with recurrent or severe, respiratory viral infections, including COVID-19, should be evaluated, especially those with evidence of irreversible inflammation and scarring (bronchiectasis, polyp disease). If this subject’s inborn error of immunity had been discovered earlier in life, some of his permanent complications may have been preventable.

In summary, we present the oldest case of an individual suffering from compound heterozygous *IFIH1* variants resulting in a phenotype of recurrent viral infections, nasal polyposis and chronic pansinusitis due to bacterial and fungal organisms, marked chronic eosinophilia, severe asthma with ground glass opacities and bronchiectasis, as well as neuropathy.

## Methods

### Human subjects

The patient enrolled in this study provided written informed consent. All methods in this study were approved by the National Human Genome Research Institute (NHGRI) central Institutional Review Board.

### Antibodies

Anti-CD3ε AF647 (HIT3a), CD19 BV421 (HIB19), CD56 AF647 (5.1H11), CD14 BV421 (HCD14), CD11c AF488 (3.9), and Human TruStain FcX were from Biolegend. Anti-MDA-5 (D74E4) was from Cell Signaling Technology. Anti-GAPDH (G-9) was from Santa Cruz Biotechnology. Fluorescently labeled anti-Mouse and anti-Rabbit secondary antibodies were from LI-COR Biosciences.

### FACS Analysis

PBMCs were isolated from whole blood by density centrifugation. Fc receptors were blocked, and samples were stained on ice for 20 min followed by three washes. Data were collected on a Cytek DxP instrument and analyzed with FlowJo (Treestar).

### Western Blots

PBMCs were stimulated overnight with the indicated concentrations of IFN-α (Cell Signaling Technology) in media comprising RPMI plus 10% FBS, 1x Pen/Strep, 10 mM HEPES, 1 mM sodium pyruvate, and 55 μM 2-mercaptoethanol. Cell pellets were lysed with RIPA buffer supplemented with HALT protease inhibitors (ThermoFisher), and insoluble material was removed by centrifugation. Reduced lysates were separated by SDS-PAGE and transferred to nitrocellulose membranes. Membranes were blocked with PBS-Tween containing 5% BSA and probed with primary and secondary antibodies. Data were collected on a Sapphire Biomolecular Imager (Azure Biosystems) and analyzed with the ‘Gels’ plugin in ImageJ.

## Data Availability

All data are included in the paper.

## Acknowledgements

We thank the patient and his family. Supported by the Jeffrey Modell Foundation (to Dr. Butte) and by an award (U01HG007703) from the National Institutes of Health (NIH) Common Fund, through the Office of Strategic Coordination and the Office of the NIH Director, to the University of California, Los Angeles (UCLA) (to Drs. Nelson, Lee, and Butte). Deidentified whole blood from healthy controls was provided through a UCLA Center for AIDS Research grant (5P30 AI028697) and the UCLA AIDS Institute.

## Declaration of interests

The authors declare no competing interests.

## Notes

### Competing Interest Statement

The authors have declared no competing interest.

